# The I-SPY COVID Adaptive Platform Trial for COVID-19 Acute Respiratory Failure: Rationale, Design and Operations

**DOI:** 10.1101/2022.05.05.22274628

**Authors:** D. Clark Files, Michael A. Matthay, Carolyn S. Calfee, Neil Aggarwal, Adam L. Asare, Jeremy R. Beitler, Paul A. Berger, Ellen L. Burnham, George Cimino, Melissa H. Coleman, Alessio Crippa, Andrea Discacciati, Sheetal Gandotra, Kevin W. Gibbs, Paul T. Henderson, Caroline A.G. Ittner, Alejandra Jauregui, Kashif T. Khan, Jonathan L. Koff, Julie Lang, Mary LaRose, Joe Levitt, Ruixiao Lu, Jeffrey D. McKeehan, Nuala J. Meyer, Derek W. Russell, Karl W. Thomas, Martin Eklund, Laura J. Esserman, Kathleen D. Liu, the ISPY COVID Adaptive Platform Trial Network

**Affiliations:** Wake Forest School of Medicine, Winston-Salem NC USA; University of California San Francisco, San Francisco, CA, USA; University of Colorado, School of Medicine, Aurora, CO, USA; Quantum Leap Healthcare Collaborative, San Francisco, CA, USA; Columbia University, Irving Medical Center, New York, NY, USA; Sanford Medical Center, Sioux Falls, SD, USA; Karolinska Institutet, Stockholm, Sweden; University of Alabama at Birmingham, Birmingham, AL, USA; University of Pennsylvania Perelman School of Medicine, Philadelphia, PA, USA; University of Southern California, Los Angeles, CA, USA; Yale School of Medicine, New Haven, CT, USA; Stanford Healthcare, Stanford, CA, USA

**Author notes:** corresponding author **Corresponding author contact information:** D. Clark Files MD, Medical Center Blvd, Wake Forest School of Medicine, Winston-Salem NC USA 27157. Collaborating authors listed in appendix. **Study sponsor contact information:** Paul Henderson PhD, Quantum Leap Healthcare Collaborative, 499 Illinois Street, San Francisco CA 94158.

## Abstract

**Introduction:** The COVID-19 pandemic brought an urgent need to discover novel effective therapeutics for patients hospitalized with severe COVID-19. The ISPY COVID trial was designed and implemented in early 2020 to evaluate investigational agents rapidly and simultaneously on a phase 2 adaptive platform. This manuscript outlines the design, rationale, implementation, and challenges of the ISPY COVID trial during the first phase of trial activity from April 2020 until December 2021.

**Methods and analysis:** The ISPY COVID Trial is a multi-center open label phase 2 platform trial in the United States designed to evaluate therapeutics that may have a large effect on improving outcomes from severe COVID-19. The ISPY COVID Trial network includes academic and community hospitals with significant geographic diversity across the country. Enrolled patients are randomized to receive one of up to four investigational agents or a control and are evaluated for a family of two primary outcomes—time to recovery and mortality. The statistical design uses a Bayesian model with “stopping” and “graduation” criteria designed to efficiently discard ineffective therapies and graduate promising agents for definitive efficacy trials. Each investigational agent arm enrolls to a maximum of 125 patients per arm and is compared to concurrent controls. As of December 2021, 11 investigational agent arms had been activated, and 8 arms were complete. Enrollment and adaptation of the trial design is ongoing.

**Ethics and dissemination:** ISPY COVID operates under a central institutional review board via Wake Forest School of Medicine IRB00066805. Data generated from this trial will be reported in peer reviewed medical journals.

**Trial registration number:** Clinicaltrials.gov registration number NCT04488081

**Strengths and limitations of this study:**

- The ISPY COVID Trial was developed in early 2020 to rapidly and simultaneously evaluate therapeutics for severe COVID-19 on an adaptive open label phase 2 platform
- The ISPY COVID Adaptive Platform Trial Network is an academic-industry partnership that includes academic and community hospitals spanning a wide geographic area across the United States
- Of December 2021, 11 investigational agent arms have been activated on the ISPY COVID Trial Platform
- The ISPY COVID Trial was designed to identify therapeutic agents with a large clinical effect for further testing in definitive efficacy trials—limitations to this approach include the risk of a type 2 error

## Introduction

Despite decades of promising pre-clinical studies and large well-organized clinical trials, the discovery of effective pharmacotherapeutics in critically ill patients has been exceedingly rare. The COVID-19 pandemic brought an unprecedented level of attention and urgency to uncover therapies for severe acute respiratory failure and acute respiratory distress syndrome (ARDS) related to COVID-19. New approaches to critical care clinical trials that may rapidly screen potentially effective therapies are urgently needed.^1^ In the early phase of the pandemic, in the winter of 2019-2020, global efforts were made to establish clinical trials and trial networks to investigate therapies for COVID-19. In this report, we describe the I-SPY COVID Trial, a phase 2 adaptive platform randomized trial in the United States designed to test and identify drugs with a large impact on improving recovery of hospitalized patients with severe COVID-19. This report focuses on the (1) trial rationale and background, (2) design, (3) operations, (4) statistical plan, (5) challenges and limitations of this approach in the context of the evolving COVID-19 pandemic during 2020-2021. This report focuses on the initial design of the ISPY COVID Trial and reflects the study protocol conduct from initial implementation in April 2020 through December 31^st^ 2021.

## Methods and Analysis

### Design: Rationale, Background, Eligibility Criteria and Exclusions

The I-SPY COVID Trial was inspired in large part by the I-SPY 2 Trial, a phase 2 adaptive platform clinical trial designed to discover novel treatments for patients with early-stage breast cancer with high risk for early recurrence.^2^ The I-SPY COVID Trial is a phase 2, multi-center, multi-arm, adaptive, open-label, randomized controlled trial designed to rapidly screen agents to identify those with potential impact to meaningfully improve outcomes for severe COVID-19 patients (**Figure 1**). Patients with confirmed COVID-19 and a modified World Health Organization COVID-19 level of ≥5 (defined here as requiring ≥ 6L/min nasal oxygen) who meet none of the exclusion criteria are eligible for the interventional and observational arms of the trial (**Table 1 & Figure 2)**. Initially, time to recovery (defined as reaching COVID-19 level ≤4 for at least two consecutive days) was the primary endpoint, and overall mortality a key secondary endpoint **(Supplemental Table 1)**. Following discussions with the Data Monitoring Committee and the Food and Drug Administration, mortality was combined with time to recovery as a family of two primary endpoints on January 15^th^ 2021.

**Table 1.**
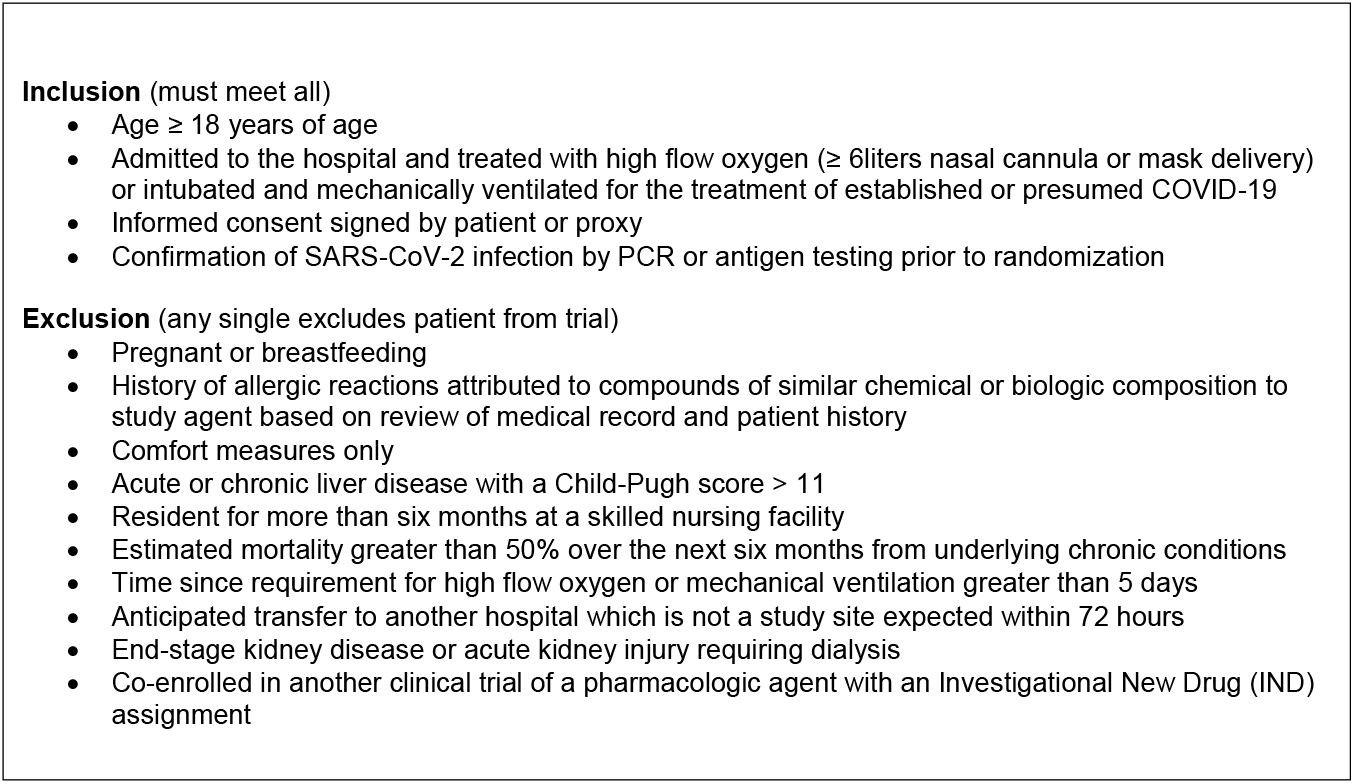
Master Inclusion Exclusion Criteria of the ISPY COVID Platform Trial

**Figure 1.**
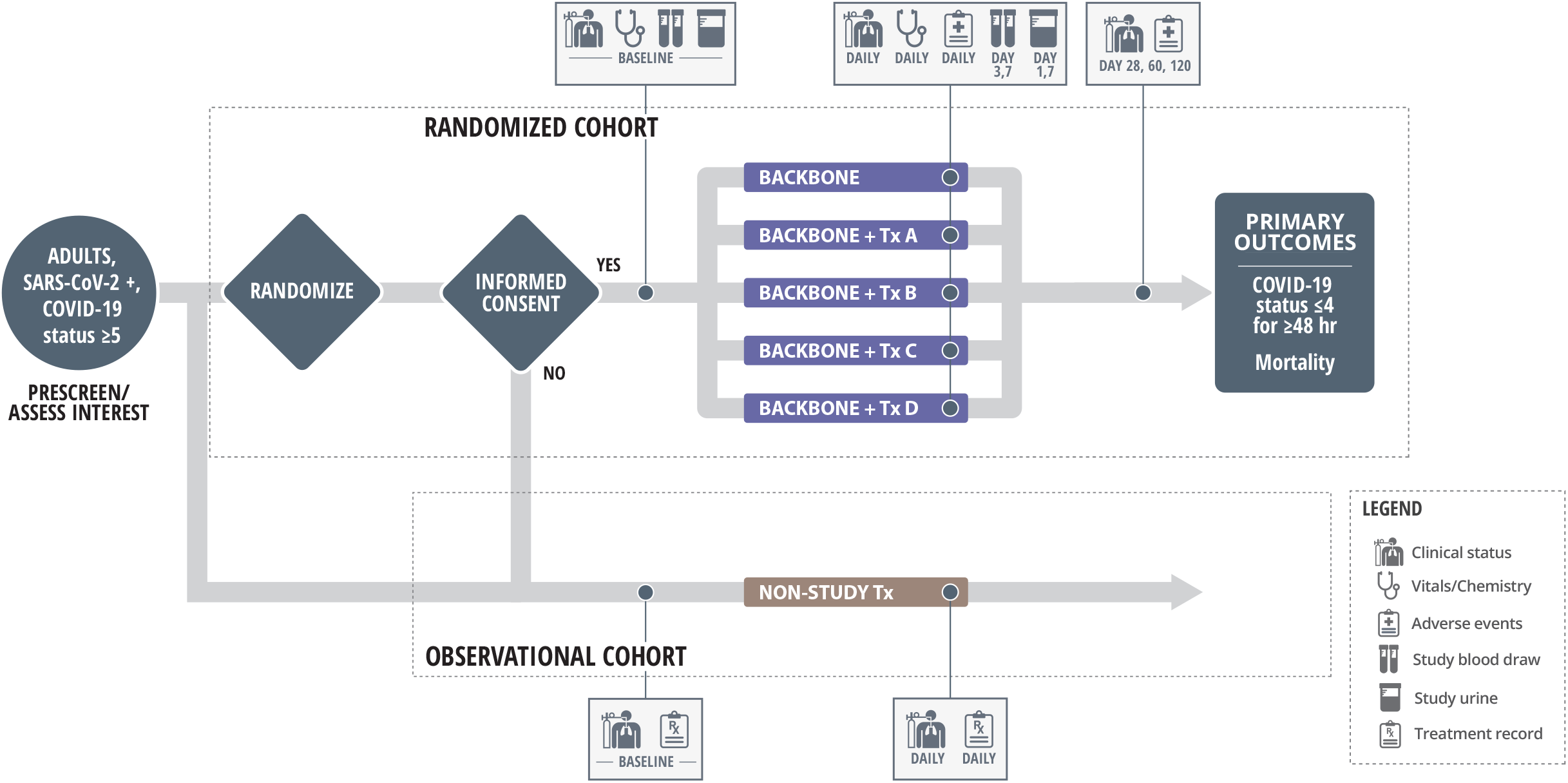
ISPY COVID Trial Design. Hospitalized COVID-19 patients who require ≥6 L/min nasal oxygen or more (COVID-19 status ≥5) are eligible for the trial. Patients or surrogates that meet master protocol inclusion/exclusion are approached for willingness to participate in the Randomized Cohort of the trial. Those that voice interest are randomized, and then approached with the agent specific consent. All patients in the Randomized Cohort receive backbone therapy with or without an additional investigational agent. Enrolled participants are followed for the assessment of the primary outcomes of resolution of severe COVID-19 and mortality. Patients that are either not approached for the Randomized Cohort of the trial or decline individual investigation agent consent are tracked in the observational cohort.

**Figure 2.**
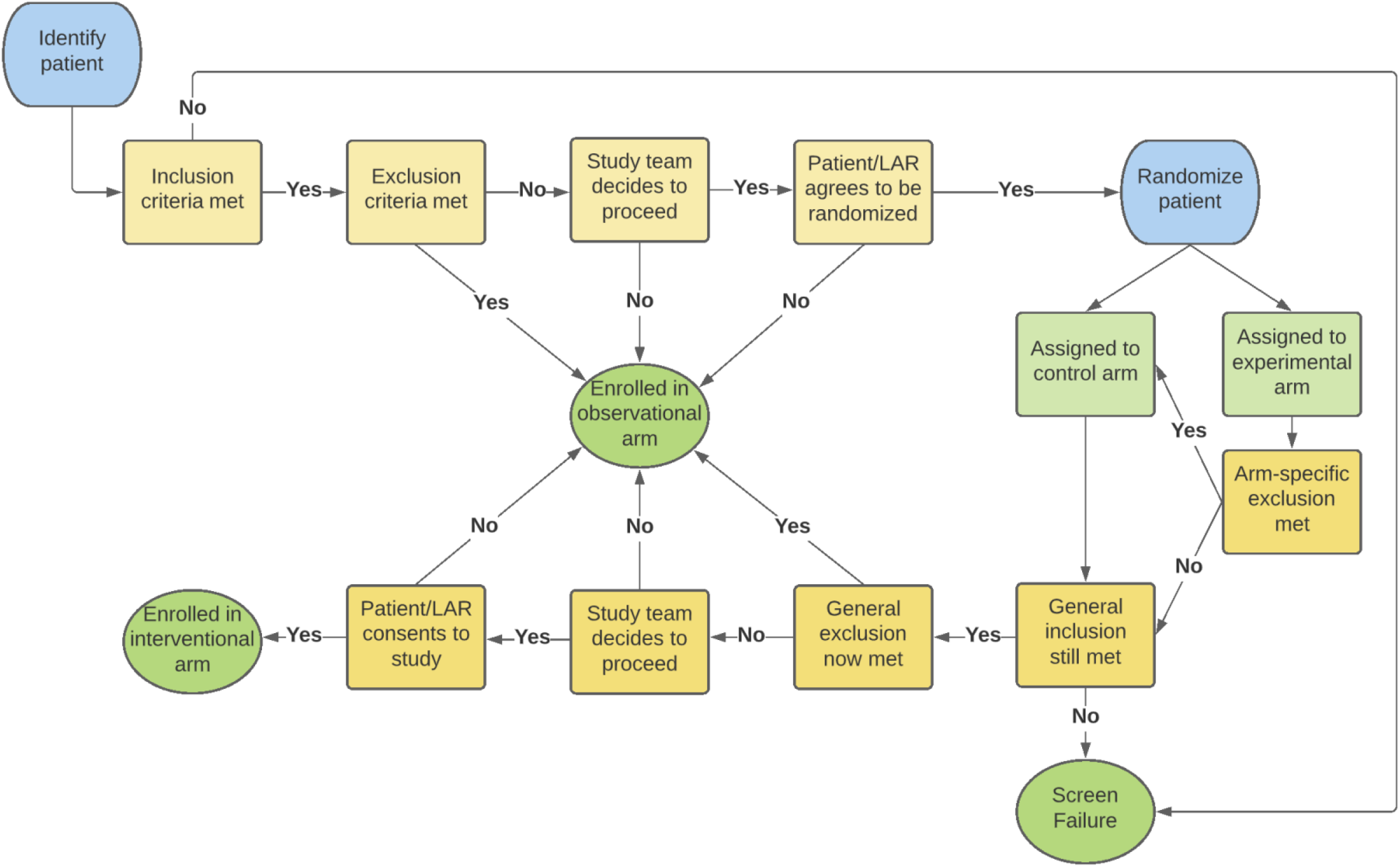
Patient Enrollment Flow into the ISPY COVID Platform Trial.

### Design: Study Backbone and Investigational Agent Selection

Patients who meet inclusion/exclusion criteria are randomized to backbone therapy alone (the control arm) or backbone therapy plus 1 of up to 4 investigational agents or agent combinations. The initial backbone treatment for the trial was assigned following review of data that emerged in mid 2020 on the use of remdesivir and dexamethasone for COVID-19 from separate clinical trials.^3 4^ During the conduct of the trial, the investigators have regularly re-evaluated the selection and dosing of dexamethasone, remdesivir and other potential therapies as new data has become available. As new therapies became available for COVID-19 (e.g. baricitinib and tocilizumab) during the conduct of the trial, the case report forms were modified to capture the clinical use of these concomitant medications.

The I-SPY COVID Agents Committee was formed to develop a process for evaluation of potential agents into the I-SPY COVID Trial. Overall agent prioritization is based on the estimated assessment of likelihood to reduce mortality and time to recovery. More specific rank criteria include a range of biologic, logistical and safety considerations as well as manufacturing supply chain capability (**Table 2**). In addition to these characteristics, practical considerations relate to the adaptive trial design, available resources for funding the testing of the therapy, and an accelerated timeline of the public health crisis. These selection criteria include a requirement for minimal drug-specific patient exclusions and low risk for drug-drug interactions in critically ill patients. High priority is assigned to the presence of a sufficient drug supply and mechanism to rapidly scale up drug production to reach a large population if treatment efficacy is established. When agents with overlapping mechanisms of action are considered, priority is assigned to one agent in that class that ranks higher in other rank criteria in order to avoid testing multiple agents within a single biologic pathway.

**Table 2.**
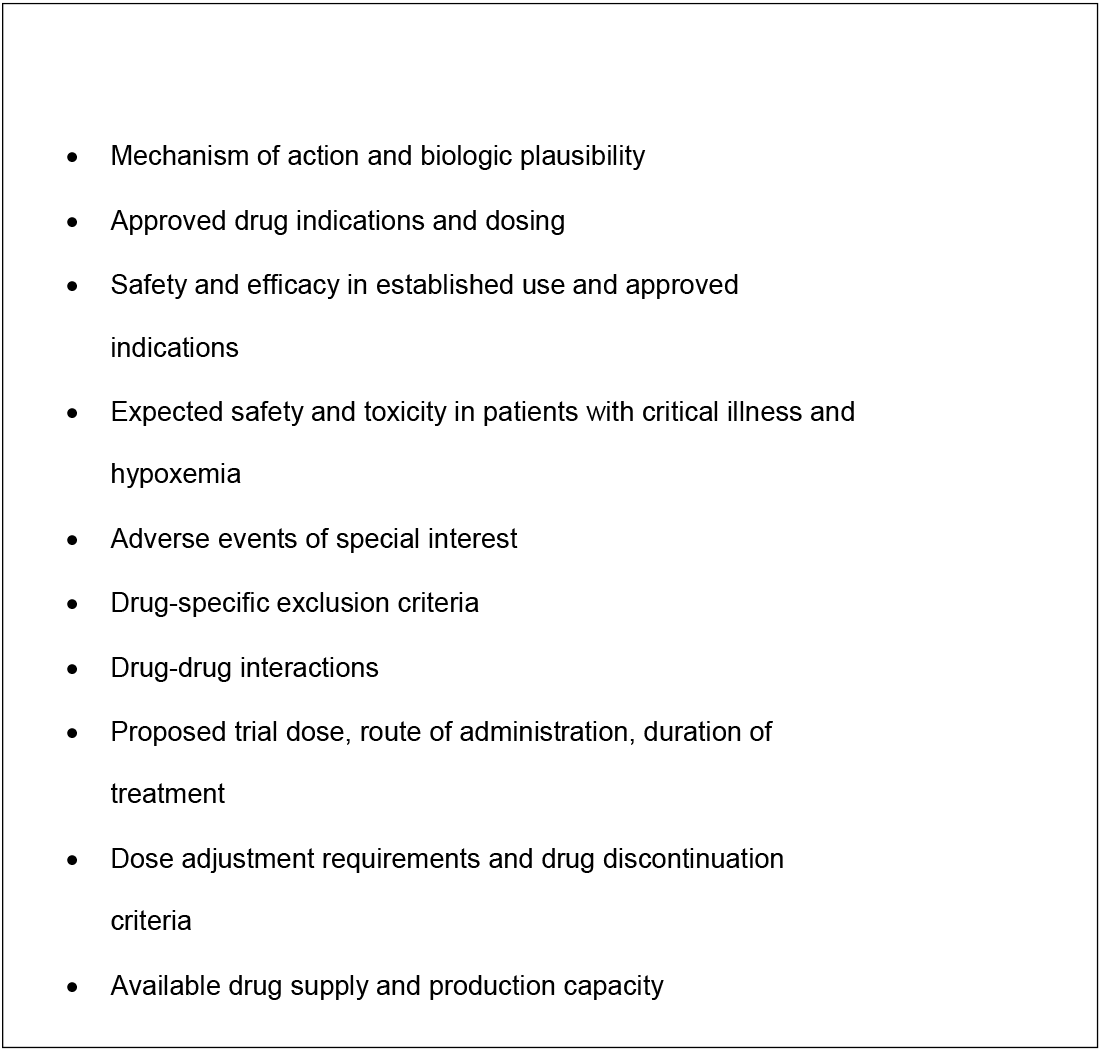
Agent Selection Criteria

Members of the Agents Committee with complementary expertise in acute lung injury, critical illness, and pharmacology evaluate each potential agent and present the higher priority candidate agents to the entire investigator group. In some instances, staff from the manufacturing company of potential agents are invited to present data directly to the investigators. However, company representatives are excluded from all follow-up deliberations and decisions on inclusion of the agent in the trial. In these subsequent discussions, pro- and con-arguments are developed and priorities for additional background research established. In twice-monthly follow-up reviews, the investigator group reaches consensus-based decisions on whether or not the agent should be included in the trial.

Throughout the agent review process, individual investigators voluntarily identify agents to support in the trial and serve as agent “chaperones” for the duration of the trial. Each agent is assigned two to three investigator chaperones. If the agent is selected for inclusion, the chaperones prepare the materials necessary for IRB approval and implementation, including agent-specific appendices to the master protocol, informed consent documents and technical guides on dosing and administration. Chaperones work closely with the clinical trial operations group to nest the agent appropriately in the context of the I-SPY Trial platform. Chaperones remain available to study site personnel throughout the treatment period to provide assistance on dosing, side effects and technical issues. The common inclusion/exclusion criteria of the platform necessitate minimizing agent-specific exclusion criteria. Therefore, significant efforts are made by chaperones and agent sponsors to streamline and eliminate as many as possible agent-specific exclusions. In instances where agent-specific exclusions become numerous, thus limiting the generalizability of the agent, the proposed agent is not selected for placement on the platform.

Investigative agents were initially identified through partnership with COVID-19 Research and Development Alliance (COVID-19 R&D),^5^ a consortium of research and development leaders in industry formed to accelerate new COVID-19 therapies and vaccines. These initially proposed agents were identified by COVID-19 R&D as having high probability of becoming successfully repurposed for COVID-19 treatment. Of the 11 agents initially identified in this process, three were selected for trial inclusion (apremilast, icatibant and cenicriviroc). Subsequent agents were brought forward for consideration through direct communication to the Agents Committee by trial investigators with expertise with a particular repurposed agent, by pharmaceutical companies responding to the opening of the trial, the United States government as well as informal contacts in academia. Between April 2020 and November 2021, over 70 individual agents were reviewed in detail, including 12 that were included in 11 trial arms (one arm was a combination of famotidine and celecoxib). Of the 8 completed arms, 5 were solicited by the pharmaceutical industry and the remainder were nominated the United States government and by trial investigators. A figure of the activated arms on the ISPY COVID trial from August 2020 through December 2021 are provided in **Supplemental Figure 1**.

### Design: Patient and Public Involvement

The primary outcomes of the ISPY COVID Trial were designed to focus on patient-centered outcomes. Both recovery of respiratory function and mortality are patient-centered endpoints. Due to the urgent nature of the pandemic, no COVID-19 patients were included in the development of the ISPY COVID Trial Protocol. The randomization followed by consent process outlined in this manuscript was developed using the process employed in the ISPY 2 Trial^6^ to reduce burden on participants and surrogates. During the development of the ISPY COVID Trial, this method was presented to the ISPY IRB Working group, which includes a patient advocate. The ISPY IRB Working group found this method to be more patient-centered and supported this method to be used in the ISPY COVID trial.

### Operations: Randomization and Consent

Up to four investigational agents can be active in I-SPY COVID at any given time. Patients are randomly assigned with equal probability to receive any of the investigational agents, while a higher proportion of participants are assigned to the backbone control arm. For instance, the ratio of randomization is 1.4:1:1:1:1 for the control arm to the four investigational arms (the ratios of control to interventional changes depending on the number of active agents in the study, with ratios 1:1 with one active agent, 1.2:1:1 with two active agents, and 1.3:1:1:1 with three active agents). Randomization is performed centrally and is stratified by site and modified WHO COVID-19 status at study enrollment.

To facilitate a patient-centered consent process, randomization is performed prior to consent. This order of the randomization and consent process has worked well in I-SPY 2 and has the advantages of avoiding a two-step consent process and simplifying patient information.^6 7^ That is, an individual patient interested in the trial only receives information about the one investigational agent that they are randomized to receive (which could also be the control arm). The disadvantage of this consent approach is that there is a risk of generating different accrual patterns across the trial arms, due to different perceived risks by participants during consent. Therefore accrual across arms must be closely monitored. Patients who do not consent to be randomized enter an observational cohort (using an IRB-approved waiver of consent mechanism), where disease outcomes and other endpoints are tracked. Patients with an agent-specific exclusion to a study drug upon randomization move into the backbone control arm. The study screening, randomization and consent process in relation to investigational drug arm, control arm or observational arm is shown in **Figure 2**.

### Rationale: The I-SPY COVID Trial-Endpoints and Open Label Design

We explicitly decided to focus on severe COVID-19, defined by clinical and physiologic criteria. This rationale is reflected in the choice of the primary endpoints of the trial. Time to recovery and mortality were thought to be important outcomes for managing the health crisis created by the global pandemic; faster time to recovery helps to increase hospital bed availability and to avoid hospital strain – a key objective during the pandemic. Mortality remains the optimal primary outcome in the field of critical care; while modification of mortality is challenging to achieve, it is still the most important patient-centered outcome.

Multiple reasons exist for the open label design for a phase 2 trial during the pandemic. Rapidly developing and executing a platform trial, whereby multiple agents are tested simultaneously, and new agents replace outgoing agents in succession, requires flexible study operations. Because different agents in the platform will have different routes of administration, dosing schedules, and durations, the use of placebos for each agent was deemed complex and unwieldy, resulting in testing fewer active agents for COVID-19. Some platform trials have used a pooled placebo concept, though this option remained impractical given the number of agents that were planned to be rapidly tested during the pandemic. While the open-label approach does potentially allow for investigator bias, a similar open label approach has also been used successfully in the RECOVERY Platform for COVID-19 in the United Kingdom.^8^

Ultimately, the open label design of the I-SPY COVID trial was deemed to be the most efficient approach to rapidly and safely evaluate novel therapeutics for severe COVID-19, with the goal that promising agents could be further tested in a closed, double blinded placebo-controlled format upon trial graduation. Lastly, we chose to include a parallel observational cohort that could be included to better understand demographics and outcomes of COVID-19 patients with rapidly changing therapeutic standards and to provide generalizability against a non-study population.

### Design: Biomarkers and Biospecimen Collections

Severe COVID-19 is characterized by an inflammatory host response to the SARS-CoV-2 virus; however, even within the group of patients with severe and critical COVID-19, there is potentially important biologic heterogeneity that may influence treatment response.^9^ The I-SPY COVID trial was designed to collect key biospecimens from enrolled patients in order to permit subsequent analyses of heterogenous treatment effect, to identify potentially important mechanisms that relate to clinical outcomes, and enable pharmacokinetic evaluations of the novel therapies being tested. Biospecimens include plasma and whole blood RNA (Days 1, 3 and 7), serum and peripheral blood mononuclear cells (PBMCs) (Days 1 and 7), urine (Days 1 and 3), and DNA (Day 1 only). Examples of the type of analyses that will be conducted include testing whether previously identified phenotypes of ARDS are relevant in COVID-19^9^, replicating innovative analyses of immunotypes within severe COVID-19,^10^ testing for anti-Type I interferon antibodies in serum from enrolled patients,^11^ and measuring plasma viral antigen levels and SARS-CoV-2 endogenous antibody levels. In addition to these exploratory biomarker analyses, biomarkers measured in clinical labs at enrolling sites such as D-dimers, CRP, and the absolute neutrophil count/absolute lymphocyte ratio are being recorded in order to test for prognostic and/or predictive enrichment value.

### Design: Statistical Analysis Plan

Bayesian survival regression models are used to model the hazard functions for the two events of interest: (i) recovery (treating death as a competing event); and (ii) overall death. We used Bayesian proportional-hazard Weibull models with weakly informative priors to model the cause-specific hazard function for recovery (treating death prior to recovery as a competing event) as a function of study arm, adjusting for baseline COVID-19 level. Similarly, Bayesian proportional-hazard Weibull models were used to model the hazard function for all-cause mortality. Importantly, concurrent controls are used so that the control group is chosen from the same population as the investigational agent group over time given the potential of changing background recovery and mortality rates in an evolving pandemic. The primary analyses are performed on the intention-to-treat (ITT) population, which includes those randomized patients who signed the informed consent.

Due to the randomization followed by consent process in this trial, the *a priori* analyses also include a *super ITT population*. This population consists of all randomized patients, regardless of whether they consented to receive the investigational agents or declined, thereby entering the observational cohort. The super ITT population will thus not be impacted by the potential effect of the randomization-consent process on the patient population in the different trial arms.

### Design: Agent Graduation and Futility Boundaries

During the course of the trial, the effect of the treatments is evaluated every two weeks by the DMC. At these evaluations, treatments may “graduate” for superiority or be dropped for futility according to the following criteria:

- If at least 50 patients have been randomized and consented to a treatment arm and the posterior probability is at least 0.975 that the cause-specific hazard ratio (csHR) for recovery (investigational agent vs. control) is greater than one OR if the posterior probability is at least 90% that the hazard ratio for overall mortality is smaller than one, the treatment is evaluated by the DMC for *graduation*.
- If at least 40 patients have been randomized and consented to a treatment arm and the posterior probability is at least 0.9 that the csHR for recovery is less than 1.5 AND the posterior probability is at least 50% that the hazard ratio for overall mortality is greater than one, a treatment is evaluated by the DMC to be dropped for futility.

If the maximum sample size of 125 participants in a treatment arm is reached, assignments to that arm will end. If an investigational agent reaches a threshold for graduation or futility, the DMC reviews the findings and make a recommendation to Study Principal Investigators (PIs) for final approval. In addition to examining hazard ratios for recovery and overall mortality, the DMC also reviews and evaluates cumulative incidence functions (**Figure 3**).

**Figure 3.**
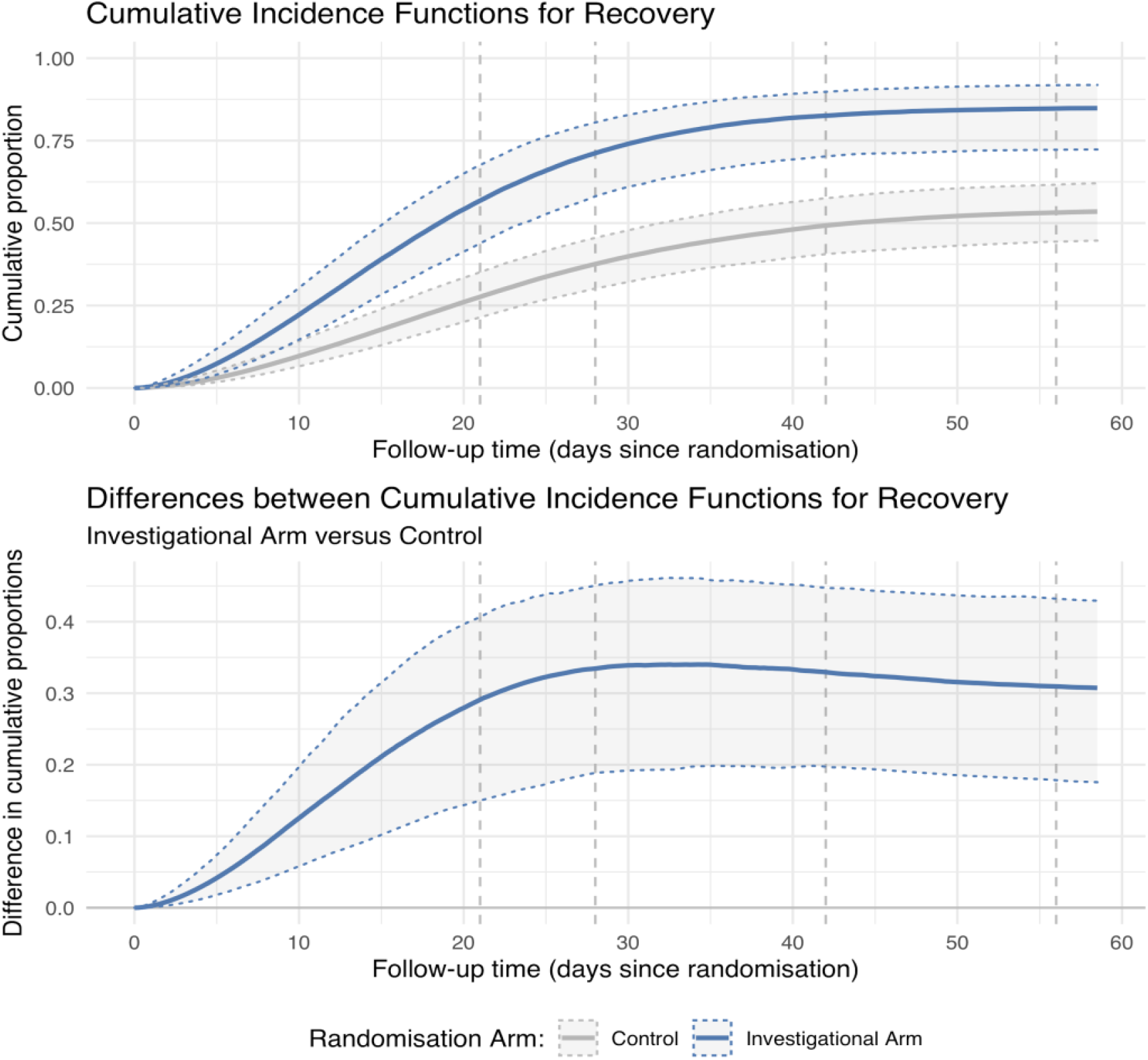
Modeling Recovery in the ISPY COVID Trial. The figure shows examples of trial data based on simulations. Top panel: Posterior cumulative incidence and survival functions (solid lines) and 95% quantile credible intervals (shaded area between dotted lines). Bottom panel: Medians (solid lines) and 95% quantile credible intervals (shaded area between dotted lines) for the posterior distribution of the difference in the Cumulative Incidence functions (Investigational Arm versus Control).

### Design: Operating Characteristics—Type 1 and Type 2 Errors

Prior to beginning the trial, study statisticians tested the trial’s operating characteristics by simulating a large number of virtual trials under multiple scenarios. The type I error rate for an individual Investigative Agent (probability to graduate a specific Investigational Agent despite no effect on reducing recovery or mortality rate) was estimated to be 4-5% when time to recovery was used a single primary endpoint. After adding overall mortality to the primary endpoint definition (to form a family of two primary endpoints, recovery and overall mortality), the type 1 error rate was estimated to up to 17% depending on simulated scenario, which was deemed acceptable for a phase 2 signal seeking trial. The power to graduate an individual agent was above 85% in scenarios where the csHR for recovery was set to 1.75. Similarly, the power for a given individual investigational agent arm was greater than 85% if the HR for overall mortality was below 0.5. Investigational Agents with more moderate effect sizes for recovery (csHRs of 1.5 or less or a HR for overall mortality of 0.7 or higher) graduated at lower rates (about 65% or less).

Overall, these simulations indicate that the current graduation and futility rules may control reasonably well the false graduation rates, while at the same time they might provide sufficient power to graduate highly effective investigational agents. However, there are important limitations to this approach including the risk of a type 2 error, due to small sample sizes, giving limited power for smaller effect sizes. The design to use 40-125 patients in each group runs the risk of discarding a potentially effective agent, ie a type 2 error. In prior phase 2 trials, wide confidence intervals illustrate the potential for a type 2 error with a restricted number of patients, as discussed by Abraham and Rubenfeld regarding a phase 2 trial of sepsis.^12^ With wide confidence intervals and small numbers of patients, it is challenging to exclude harm or benefit. Another example is the Brower trial of lung protective ventilation in 52 patients which showed no benefit,^13^ but then the properly powered ARMA trial with 861 patients showed a major reduction in mortality.^14^

The I-SPY COVID Trial was designed early in the pandemic, and given the singular etiology of lung injury in patients with severe COVID-19, many investigators anticipated that ARDS from COVID-19 would exhibit less heterogeneity than “traditional” ARDS.^15^ While it remains unclear whether severe COVID-19 ARDS exhibits the degree of heterogeneity of ARDS in the pre-COVID-19 era,^9 16^ further biologic and clinical phenotyping may be necessary to find effective targeted therapies in a screening trial of this size and potentially re-evaluating the number of patients needed to evaluate candidate agents.

### Operations: Real Time Data Entry and Reporting

A minimal set of key clinical and research outcome data elements were defined as part of the daily checklist for enrolled participants (**Figure 4)**. The “checklist” was implemented in OpenClinica (Waltham, MA) Electronic Data Capture (EDC) system to support data entry for subjects randomized to investigational agent arms or the observational cohort. Daily eCRFs are completed from enrollment until discharge, with additional follow up consisting of electronic Patient Reported Outcome (ePRO) survey questionnaires using HealthMeasures PROMIS© and Patient-Reported Outcomes version of the Common Terminology Criteria for Adverse Events (PRO-CTCAE™) validated instruments at day 28, 60 and 120.

**Figure 4.**
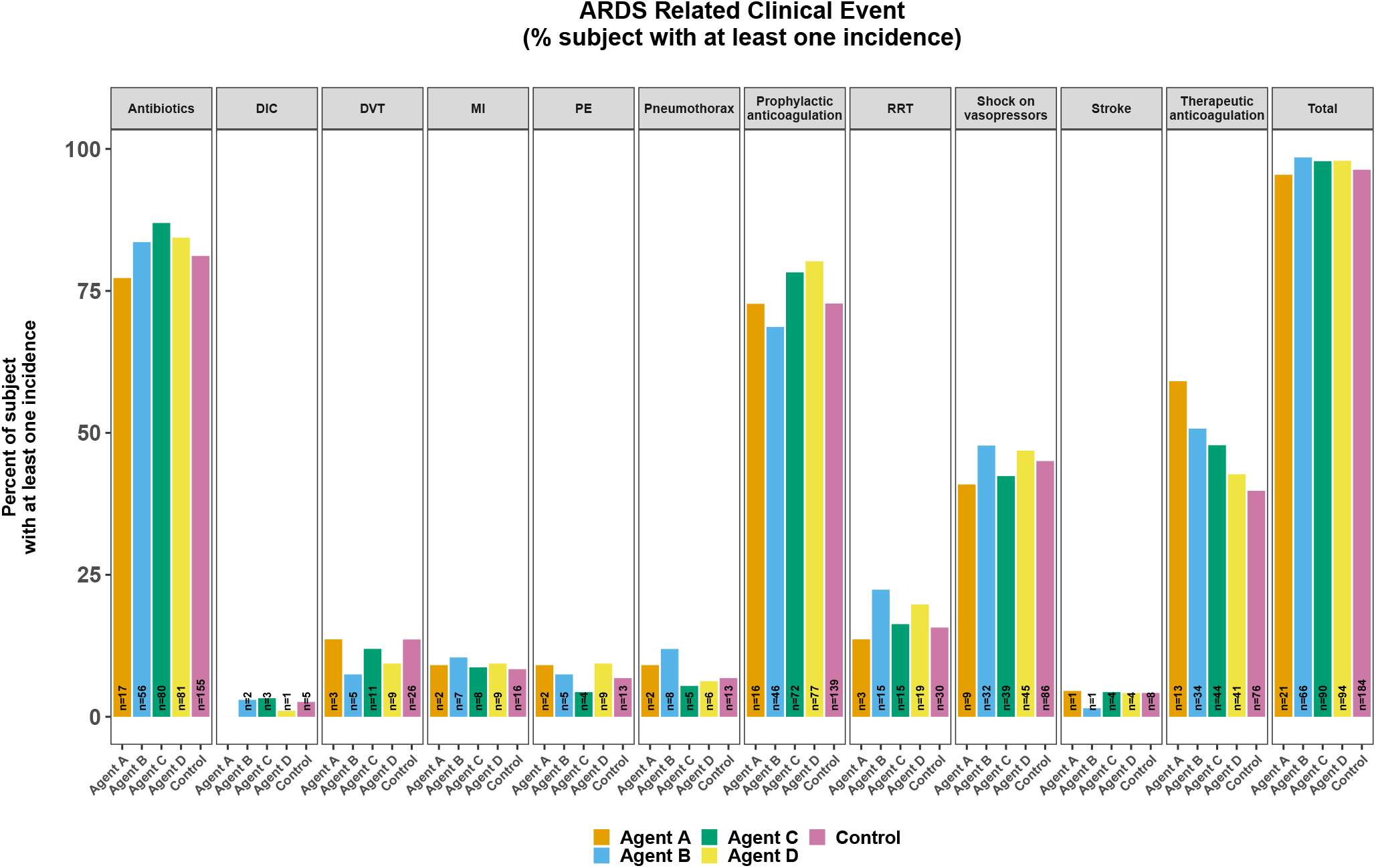
Daily Clinically Important Events Systematically Captured in Participants. Daily forms are completed in ISPY participants relating to clinically important events occurring in the context of severe COVID-19. These events are then systematically reported across arms of the study for safety reports to the Data Monitoring Committee.

Despite streamlining the data collection in I-SPY COVID, the pandemic strains on clinical and research staff sometimes led to delays in data entry that were addressed on a site-specific basis using remote staffing models (see “Novel Implementation Challenges” section below). Limitations to a streamlined dataset include reduced granularity of exploring important physiologic variables such as oxygenation index or plateau pressures, or the Fi0_2_ or flow rates on patients enrolled being treated with high-flow nasal oxygen, which have important clinical implications for patients with acute respiratory failure from severe lung injuries.

### Operations: Safety

Clinical trials in critically ill patients share unique challenges for safety monitoring. Specifically, study participants often possess significant comorbidities (i.e., chronic organ dysfunction, immune suppression, and malignancy), receive numerous concomitant medications with potential for interactions, and experience marked derangements in baseline physiology (i.e., vital sign and laboratory abnormalities) relative to populations in which potential investigational agents have previously been studied. These challenges are amplified for COVID-19 patients where multisystem organ involvement is common but accurate baseline rates of complications such as stroke, thrombosis, and cardiac dysfunction are yet to be established, and may vary over time. Additionally, the usual care of these patients is a rapidly moving target. This topic is particularly challenging for phase 2 trials in which there may be limited clinical experience for investigational agents and even FDA-approved medications may be used at higher doses and in combinations not previously studied. The open-label and shared control structure of the I-SPY COVID platform trial is designed to allow for monitoring for known side effects and rapid testing of multiple agents, but also creates potential for bias in adverse event reporting due to differential scrutiny applied to study arms (i.e., monitoring for secondary infections in patients randomized to an immunomodulatory agent or kidney injury for a potentially nephrotoxic agent relative to shared controls). The I-SPY COVID platform has used several strategies to overcome these challenges.

First, laboratory assessment of vital signs and organ dysfunction are collected daily and clinically important events (i.e., pulmonary embolism, deep venous thrombosis, and kidney injury) are systemically collected on daily report forms. This approach allows for systematic comparisons across therapy arms of event rates and organ failures rather than relying on investigator recognition and capture of these events (**Figures 4 and 5**). Second, I-SPY COVID has constituted a Safety Working Group (SWG) to provide an additional layer of structured safety monitoring across all trials within the platform. The SWG provides real-time monitoring of adverse events and provides guidance to the DMC on adverse event reporting. The SWG is led by two critical care physicians not otherwise part of the clinical trial and also includes the drug chaperones, study principal investigators, and operations committee chairs. This group meets on a regular basis to review adverse events and to determine potential attribution to COVID-19 and/or to investigational agents. The SWG chairs also provide external review of severe adverse events and other safety events that might require expedited reporting. Finally, the drug chaperones serve as internal content experts for a given agent and are available to investigators on a 24/7 basis to review potential events related to the investigational agent regardless of expectedness. Reporting of potential events is encouraged so that these can be formally reviewed by the Safety Working Group. Every death, AE, AESIs and SAE is reviewed by the Safety Working Group to evaluate if the adverse event is expected for the investigational agent, if expected with COVID-19 or ARDS, and likelihood of being caused by the investigational agent. Adverse event reporting is configured using the Shiny R Studio. Lastly, site conduct is audited by Quantum Leap Healthcare Collaborative and trial conduct by other study sponsors.

**Figure 5.**
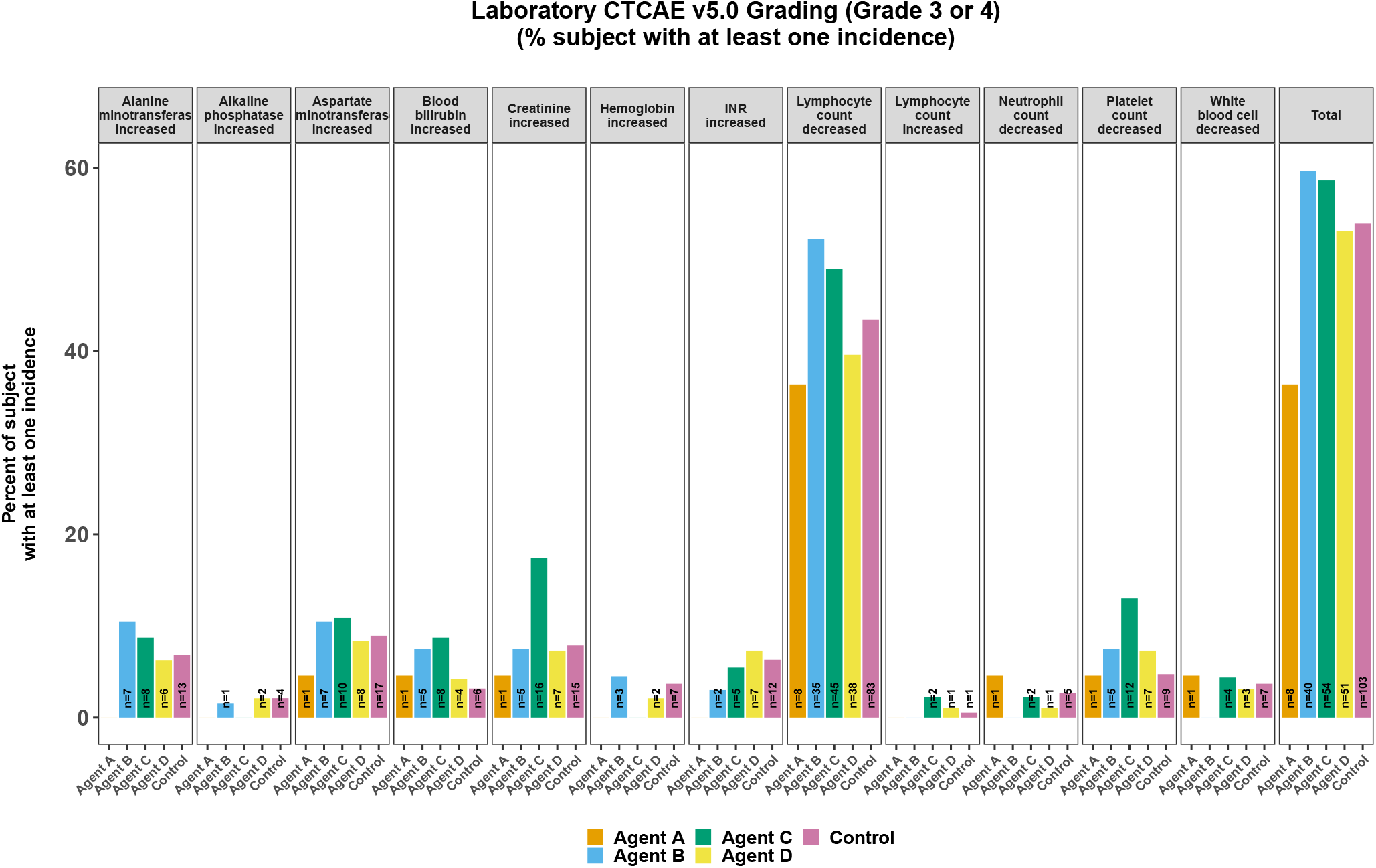
Severe Laboratory Abnormalities Systematically Captured in Participants. Potentially significant daily laboratory data entered by study coordinators is systematically captured, graded and reported back to the Data Monitoring Committee to evaluate safety. In this simulated example, Agent C appears to be associated with increased creatinine, suggesting renal toxicity.

### Operations: Pharmacy

The investigational drug services (IDS) pharmacy plays a pivotal role in the ability to deploy agents safely and rapidly into a continuously running adaptive platform trial, such as I-SPY COVID. Careful planning and strong communication are paramount, but several additional key issues must be addressed to accomplish this efficiently. First, participating sites typically designate one lead and at least one back-up pharmacist at each site. Second, a consistent workflow (standard operating procedure) for the implementation of each new agent is established. This workflow includes centralized delivery of drug and trial arm-related information, personnel training documentation procedures, and drug storage provisions. Third, IDS pharmacists implement drug ordersets into the clinical workflows at each site.

Given the rapidity with which new agents enter the trial, pharmacy lead time and preparedness is important. Adequate lead-time is required to allow for the preparations mentioned above to be safely and smoothly executed. As such, for a trial of this nature in which study arms may drop out without warning and new arms quickly advance in the priority list, the coordinating center must ensure that all affiliated pharmacies are prepared for future agents before they are added to the trial.

For study sites that do not already have an Investigational Drug Services (IDS) Pharmacy, the aforementioned recommendations are even more critical. In addition to those, it is also vital to identify a pharmacist (and back-up) committed to overseeing the pharmaceutical aspects of the study at both the community partner site and the supervising site. An electronic drug accountability system that meets FDA requirements for investigational drugs is necessary to allow for ease of monitoring for drug usage and storage. A remote (or in-person) visit from the partnering IDS pharmacy can help ensure that drug storage conditions are appropriate and that evaluation as well as documentation (e.g., of temperature controls) can be appropriately conducted and any gaps in training, equipment, space, or hours of site pharmacy coverage or expertise can be identified and addressed.

### Additional Considerations: No established network or funding, contracting

To rapidly meet the short timeline of the pandemic, I-SPY COVID leveraged the efficiency and infrastructure of the I-SPY 2 Trial, partnering with the not-for-profit sponsor, Quantum Leap Healthcare Collaborative. One of the solutions to drive efficiency was to first activate sites that already had existing I SPY 2 contracts. As well, we used the same contract for every site. All pharma companies also agree to a single contract. The prior experience with many of the companies sped the process of working with companies. As well, the initial partnership with the COVID R&D consortium established an example for how companies could work collaboratively and quickly in a pandemic. Some sites (primarily those not familiar with I SPY 2) had longer delays with contracting and site activation. Establishing a central IRB was essential for this process. The ISPY COVID trial is composed primarily of academic medical centers with experience conducting critical care clinical trials. However, the group felt it important to also include community sites, where most patients across the country receive care for COVID-19. The group used hybrid approaches to activate and support community sites without significant research infrastructure using different site involvement paradigms (**Table 3**).

**Table 3.** Community Site Infrastructure Paradigms in ISPY COVID

| Types of hospitals | Status | Pharmacy | Coordinators | Compensation | Geographic |
| --- | --- | --- | --- | --- | --- |
| **Academic or Community Centers with Research Capacity and Experience** | I-SPY COVID site | Investigational Pharmacy | I-SPY COVID site | Full | NA |
| **Academic or Community Centers with ICU care but Limited Research Infrastructure** | Partnership with one of the I SPY COVID sites | Clinical Pharmacist trained by Investigational Pharmacy Partner | In partnership with one of the I-SPY COVID sites | Shared | NO |
| **Community Hospitals without Sufficient ICU Care for Severe COVID-19** | Transfer patients to nearby I SPY COVID site | Transfer to I-SPY Site | Partner with I-SPY site. Work to identify eligible patients and coordinate transfer | Shared | YES |

### Additional Considerations: Novel Implementation Challenges During the Pandemic

The alarming pace of the COVID-19 pandemic has placed substantial time pressure on therapeutic trials.^17^ Hospitals surging with COVID-19 patients have potential to be the largest contributors to trials. Yet, frequently, they are also the most resource-strained. Shortages of such basic necessities as personal protective equipment (PPE), medications, and personnel have occurred often.^18^ In many hospitals, investigators and clinical research staff with relevant skills were reassigned to clinical duties. Several hospitals and universities, concerned for anticipated revenue losses, instituted broad hiring freezes or required new arduous administrative approvals to hire staff, which in some cases unintentionally impeded timely scaling up of trial personnel.

To address these issues, I-SPY COVID took several innovative steps. The trial secured PPE for shipment to sites as needed, helping ensure trial procedures would not deplete PPE in hospitals facing extreme scarcity. To address potential medication shortages, the trial committed to providing an independent supply of backbone therapies received by all patients, which included remdesivir at the time of initial trial design.

Staffing shortages have been addressed in part by partnering with a healthcare staffing company to hire off-site data entry specialists, freeing on-site staff to focus on patient accrual and in-person study procedures during surges. This “rapid response” staffing model has the added benefit of being able to reassign data entry specialists already familiar with the protocol and case report forms to new locations as surges wax and wane among sites over time. To overcome in-person research staff shortages, high-enrolling sites have engaged qualified, approved clinical staff in the recruitment and consent process—embracing a shared mission of expanding access to promising therapeutics and accelerating discovery, overcoming the conventional clinical/research divide.

Each of the above sections includes comments on the challenges for the design and conduct of this I-SPY COVID platform trial during the COVID-19 pandemic. The investigators will consider future modifications to the protocol, including moving consent prior to randomization, decreasing the number of agents to be tested at the same time, reviewing the graduation and futility criteria, and the sample size as well as increasing the granularity of baseline systemic and respiratory data collection. The strengths and weaknesses of the open-label versus a placebo-controlled design will also be evaluated. We now know that many factors contribute to heterogeneity of COVID-19 disease severity in hospitalized patients so further consideration of ways to enhance the database but maintain reasonable feasibility and efficiency will be evaluated.

## Ethics and Dissemination

The trial procedures and protocols are regulated under a central IRB structure at the Wake Forest School of Medicine. All patients (or designated surrogate) entering the portion of the trial that receive an investigational agent undergo patient level consent by study staff and/or study investigators. Protocol revisions are announced at weekly investigator and coordinator meetings and are submitted to the FDA and IRB. No patient level data will be released, and all personal information will remain confidential and de-identified. Results of the agents completing the ISPY COVID Trial will be reported in press releases, scientific abstracts and manuscripts.

## Contributorship Statement

DCF, MAM, CSC, NA, JRB, PAB, ELB, GC, SG, KWG, PTH, KTK, JLK, JL, NJM, DWR, KWT, ME, LJE, KDL contributed to the study protocol design and drafting of the manuscript. ALA, MHC, JL, PTH, CAGI, AJ, ML, JDM, DCF, MAM, CSC, LJE, KDL contributed to the study operations. AC, AD, RL, ME, LE contributed to the statistical design. All authors reviewed and provided final edits.

## Data Sharing Statement

Data generated from the ISPY COVID Trial will be made available in peer reviewed journals.

---

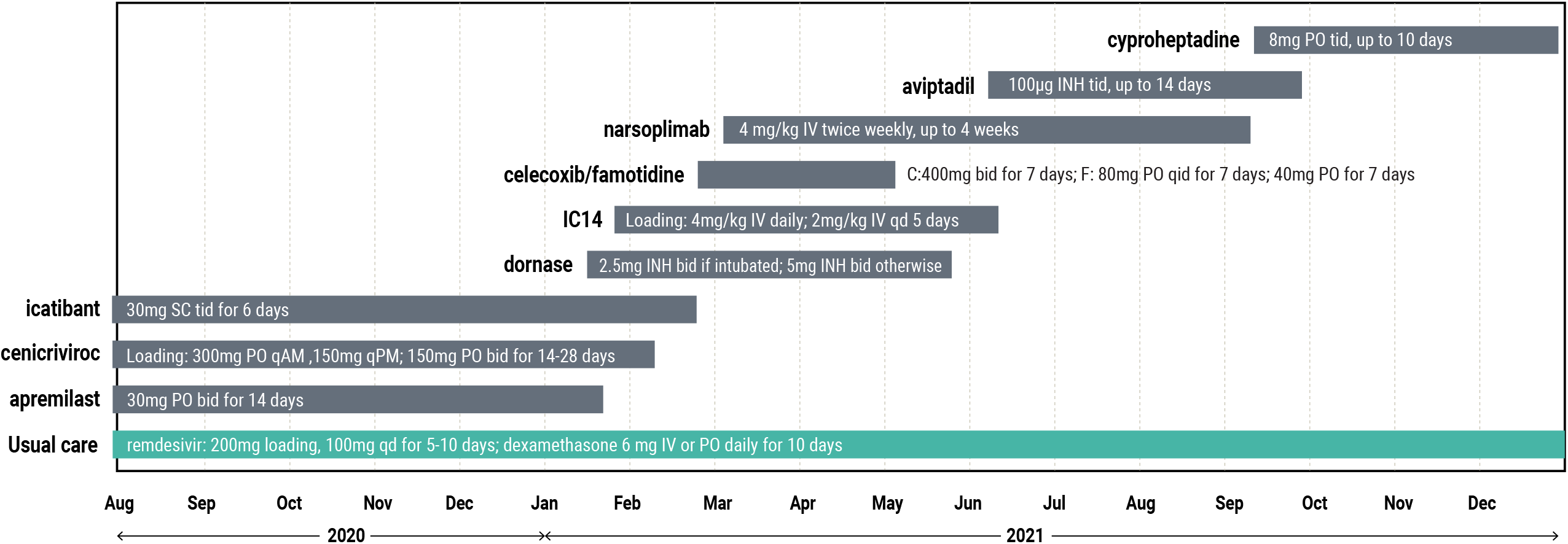

## References

1. Goligher EC, Zampieri F, Calfee CS, et al. A manifesto for the future of ICU trials. Critical care 2020;24(1):686. doi: 10.1186/s13054-020-03393-5 [published Online First: 2020/12/11]

2. Esserman LJ, Woodcock J. Accelerating identification and regulatory approval of investigational cancer drugs. Jama 2011;306(23):2608–9. doi: 10.1001/jama.2011.1837 [published Online First: 2011/12/22]

3. Beigel JH, Tomashek KM, Dodd LE, et al. Remdesivir for the Treatment of Covid-19 - Final Report. The New England journal of medicine 2020;383(19):1813–26. doi: 10.1056/NEJMoa2007764 [published Online First: 2020/05/24]

4. Group RC, Horby P, Lim WS, et al. Dexamethasone in Hospitalized Patients with Covid-19. The New England journal of medicine 2021;384(8):693–704. doi: 10.1056/NEJMoa2021436 [published Online First: 2020/07/18]

5. https://www.covidrdalliance.com.

6. Barker AD, Sigman CC, Kelloff GJ, et al. I-SPY 2: an adaptive breast cancer trial design in the setting of neoadjuvant chemotherapy. Clin Pharmacol Ther 2009;86(1):97–100. doi: 10.1038/clpt.2009.68 [published Online First: 2009/05/15]

7. Wang H, Yee D. I-SPY 2: a Neoadjuvant Adaptive Clinical Trial Designed to Improve Outcomes in High-Risk Breast Cancer. Curr Breast Cancer Rep 2019;11(4):303–10. doi: 10.1007/s12609-019-00334-2 [published Online First: 2020/12/15]

8. Normand ST. The RECOVERY Platform. The New England journal of medicine 2021;384(8):757–58. doi: 10.1056/NEJMe2025674 [published Online First: 2020/07/25]

9. Sinha P, Calfee CS, Cherian S, et al. Prevalence of phenotypes of acute respiratory distress syndrome in critically ill patients with COVID-19: a prospective observational study. The Lancet Respiratory medicine 2020;8(12):1209–18. doi: 10.1016/s2213-2600(20)30366-0 [published Online First: 2020/08/31]

10. Mathew D, Giles JR, Baxter AE, et al. Deep immune profiling of COVID-19 patients reveals distinct immunotypes with therapeutic implications. Science 2020;369(6508) doi: 10.1126/science.abc8511 [published Online First: 2020/07/17]

11. Combes AJ, Courau T, Kuhn NF, et al. Global absence and targeting of protective immune states in severe COVID-19. Nature 2021;591(7848):124–30. doi: 10.1038/s41586-021-03234-7 [published Online First: 2021/01/26]

12. Rubenfeld GD, Abraham E. When is a negative phase II trial truly negative? American journal of respiratory and critical care medicine 2008;178(6):554–5. doi: 10.1164/rccm.200807-1136ED [published Online First: 2008/08/30]

13. Brower RG, Shanholtz CB, Fessler HE, et al. Prospective, randomized, controlled clinical trial comparing traditional versus reduced tidal volume ventilation in acute respiratory distress syndrome patients. Critical care medicine 1999;27(8):1492–8. doi: 10.1097/00003246-199908000-00015 [published Online First: 1999/09/02]

14. Ventilation with Lower Tidal Volumes as Compared with Traditional Tidal Volumes for Acute Lung Injury and the Acute Respiratory Distress Syndrome. New England Journal of Medicine 2000;342(18):1301–08. doi: doi:10.1056/NEJM200005043421801

15. Juschten J, Tuinman PR, Guo T, et al. Between-trial heterogeneity in ARDS research. Intensive care medicine 2021;47(4):422–34. doi: 10.1007/s00134-021-06370-w [published Online First: 2021/03/14]

16. Grasselli G, Tonetti T, Protti A, et al. Pathophysiology of COVID-19-associated acute respiratory distress syndrome: a multicentre prospective observational study. The Lancet Respiratory medicine 2020;8(12):1201–08. doi: 10.1016/s2213-2600(20)30370-2 [published Online First: 2020/08/31]

17. Lane HC, Fauci AS. Research in the Context of a Pandemic. The New England journal of medicine 2021;384(8):755–57. doi: 10.1056/NEJMe2024638 [published Online First: 2020/07/18]

18. Ranney ML, Griffeth V, Jha AK. Critical Supply Shortages - The Need for Ventilators and Personal Protective Equipment during the Covid-19 Pandemic. The New England journal of medicine 2020;382(18):e41. doi: 10.1056/NEJMp2006141 [published Online First: 2020/03/27]

