## Supplemental Data for "The I-SPY COVID Adaptive Platform Trial for COVID-19 Acute Respiratory Failure: Rationale, Design and Operations"

Data Supplement

Supplemental Table 1. Modified WHO Ordinal Scale

| **Score** | **Description** |
| --- | --- |
| 0 | No clinical or virologic evidence of infection |
| 1 | Not hospitalized, no limitation on activities |
| 2 | Not hospitalized, limitation on activities |
| 3 | Hospitalized, not requiring supplemental oxygen |
| 4 | Hospitalized, requiring supplemental oxygen (<6 liters/minute by nasal cannula or mask) |
| 5 | Hospitalized, on non-invasive ventilation or high-flow oxygen devices (≥6 liters/minute by nasal cannula or mask) |
| 6 | Hospitalized, on invasive mechanical ventilation |
| 7 | Hospitalized, ventilation plus additional organ support-vasopressors, renal replacement therapy or extracorporeal membrane oxygenation |
| 8 | Death |

**Supplemental Figure 1 Legend**. Activated agent arms and doses in the ISPY COVID trial from August 2020 through December 2021. The figure demonstrates the ability of the open label platform design to simultaneously evaluate up to four active agent arms at a given time. One agent that was briefly active, razuprotafib is not included, as monitoring requirements for that study drug administration were challenging during the height of the pandemic, and it was pulled from the study.
